# Univariable and multivariable Mendelian randomization study identified the key role of gut microbiota in immunotherapeutic toxicity

**DOI:** 10.1101/2023.07.24.23292742

**Authors:** Baike Liu, Zheran Liu, Tianxiang Jiang, Xiangshuai Gu, Xiaonan Yin, Zhaolun Cai, Xiaoqiao Zou, Lei Dai, Bo Zhang

## Abstract

**Background:** In cancer patients receiving immune checkpoint inhibitors (ICIs), there is emerging evidence suggesting a correlation between gut microbiota and immune-related adverse events (irAEs). However, the exact roles of gut microbiota and the causal associations are yet to be clarified.

**Methods:** To investigate this, we first conducted a univariable bi-directional two-sample Mendelian randomization (MR) analysis. Instrumental variables (IVs) for gut microbiota were retrieved from the MiBioGen consortium (18,340 participants). GWAS summary data for irAEs were gathered from an ICIs-treated cohort with 1,751 cancer patients. Various MR analysis methods, including Inverse variance weighted (IVW), MR PRESSO, maximum likelihood (ML), weighted median, weighted mode, and cML-MA-BIC were used. Furthermore, multivariable MR (MVMR) analysis was performed to account for possible influencing instrumental variables.

**Results:** Our analysis identified fourteen gut bacterial taxa that were causally associated with irAEs. Notably, *Lachnospiraceae* was strongly associated with an increased risk of both high-grade and all-grade irAEs, even after accounting for the effect of BMI in the MVMR analysis. *Akkermansia*, *Verrucomicrobiaceae*, and *Anaerostipes* were found to exert protective roles in high-grade irAEs. However, *Ruminiclostridium6*, *Coprococcus3*, *Collinsella*, and *Eubacterium (fissicatena group)* were associated with a higher risk of developing high-grade irAEs. *RuminococcaceaeUCG004*, and *DefluviitaleaceaeUCG011* were protective against all-grade irAEs, whereas *Porphyromonadaceae*, *Roseburia*, *Eubacterium (brachy group)*, and *Peptococcus* were associated with an increased risk of all-grade irAEs.

**Conclusion:** Our analysis highlights a strong causal association between *Lachnospiraceae* and irAEs, along with some other gut microbial taxa. These findings provide potential modifiable targets for managing irAEs and warrant further investigation.

## Introduction

Applications of immune checkpoint inhibitors (ICIs), especially those targeting CTLA-4 and PD-1/PD-L1, have revolutionized the treatment of various aggressive cancers (1). By blocking inhibitory signaling pathways and reinvigorating the natural anti-tumor immune response, these inhibitors have significantly prolonged the lives of numerous cancer patients (2–5). However, due to the inhibition of the systemic brake of immune activation, ICIs can cause off-target effects resulting in immune-mediated toxicities to organs and non-malignant tissues. This newly emerging registry of iatrogenic effects, known as immune-related adverse events (irAEs), usually resemble autoimmune disorders, such as colitis, dermatitis, and thyroiditis (6). Although the majority of irAEs manifest in a mild manner, still, up to 55% of patients develop serious irAEs in combined therapy (anti-CTLA-4 and anti-PD-1) (7). Notably, serious irAEs pose significant risks to patients’ well-being and may result in morbidity and mortality, not only due to the adverse event itself but also due to the need to suspend or terminate ICIs therapy and the potential impairment of the ICIs-induced immune response while using immunosuppressants (e.g. corticosteroids) (8–10). Therefore, effective management of irAEs is critical to optimize the safety and efficacy of ICIs therapy.

The precise mechanisms underlying irAEs are not fully understood, but emerging evidence indicates that the gut microbiota, a complex and dynamic system of microorganisms colonizing the intestinal tract, may play a crucial role in the regulation of irAEs. Simpson et al. found a reduced alpha-diversity of intestinal microbiota in patients who developed severe irAEs (11). Furthermore, antibiotics commonly prescribed prophylactically to hospitalized patients have been shown to increase the risk of ICIs therapy-related irAEs that are not limited to the gastrointestinal tract (12–14). The gut microbiota closely interacts with the host immune system and has been implicated in the regulation of various autoimmune and inflammatory disorders (15,16). However, consensus on the core microbial drivers or protective microbes of irAEs is still lacking, due to inconsistent findings reported in previous studies (11,17–19). The discrepancies among previous studies may be attributed to limited sample sizes and susceptibility to confounding factors such as age, diet, and medication usage in observational designs (11,20).

Mendelian randomization (MR), initially described by Katan in 1986 (21), is a novel method for inferring causal associations between modifiable risk factors and health outcomes using genetic variations as instrumental variables (IVs) (22). MR effectively addresses the limitations of confounding and measurement errors that often exist in observational studies, as the direction of causation is from the genetic polymorphism to the trait of interest, not vice versa (23). Therefore, we aim to utilize MR, an increasingly popular method in drug discovery and epidemiology (24,25), to investigate the potential association between the gut microbiota and irAEs, providing further evidence for the management of irAEs by manipulating human gut microbiota.

## 2. Methods

### Study design and data source

An overview of the study design was illustrated in **Figure 1**. In general, we first performed univariable bi-directional two-sample MR analysis, which utilizes single nucleotide polymorphisms (SNPs) from summary-level data as proxies for the risk factor under investigation. Then, several sensitivity analyses and multivariable MR analysis (MVMR) was conducted to further increase the robustness of our study. To ensure the validity of the MR results, three assumptions needed to be satisfied, as illustrated in **Figure S1** (22):

(1) **Relevance assumption:** The genetic variants should demonstrate a strong association with the exposure;
(2) **Independence assumption:** The genetic variants should not be associated with any confounders that could affect the relationship between the exposure and outcome;
(3) **Exclusion restriction assumption:** The genetic variants should not have an independent effect on the outcome aside from their impact through the exposure.

**Figure 1.**
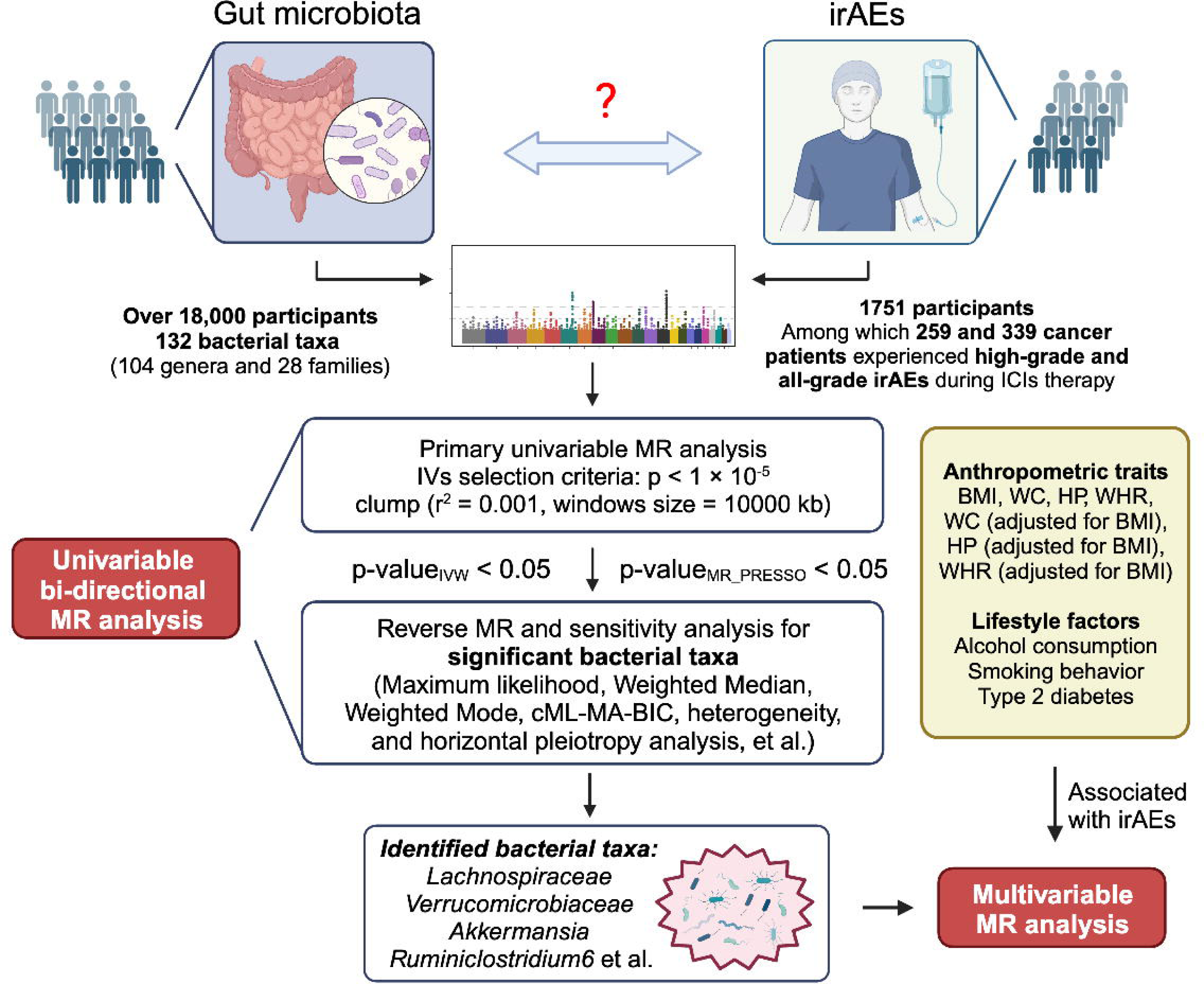
Overview of the study design. Initially, IVs were selected from the summary GWAS data of the gut microbiota and irAEs. Subsequently, by employing thresholds of p-value (IVW) < 0.05 and p-value (MR PRESSO) < 0.05, the identified gut microbiota that exhibited statistically significant associations were taken into further analysis. irAEs = immune-related adverse events; ICIs = immune checkpoint inhibitors; BMI = body mass index; WC = Waist circumference; HP = Hip circumference; WHR = Waist-to-hip ratio. (Created with BioRender.com).

This study is based on publicly available GWAS summary statistics and ethical approvals were acquired by the original studies.

### Gut microbiota

Genetic variations associated with the composition of gut microbiota were derived from the most comprehensive genome-wide meta-analysis conducted to date by the MiBioGen consortium (26). This study included a total of 18,340 individuals from 24 cohorts worldwide, mainly of European descent (n = 13,266). Fecal DNA was extracted, and targeted sequencing of variable regions in the bacterial 16S rRNA gene (V4, V3-V4, and V1-V2 regions) was performed to profile the gut bacterial composition. To account for sequencing depth differences across cohorts, all cohorts were rarefied to 10,000 reads per sample. Taxonomic classification was performed using direct taxonomic binning method (26). Following quality control, imputation, and post-imputation filtering procedures, gut bacterial taxa observed in over 10% of samples were included in the microbiota quantitative trait loci (mbQTL) mapping (26). This allowed us to identify host genetic variants associated with the relative abundance of bacterial taxa. Further details on microbial data processing can be found in the original study. Genus-level and family-level taxa were included in our analysis, resulting in a total of 131 genus-level and 35 family-level taxa.

### irAEs

Summary-level data of irAEs was obtained from a recent GWAS conducted in the Dana-Farber Cancer Institute (DFCI) cohort (27). The study included 1,751 cancer patients of European ancestry who underwent ICIs treatments between 2013 and 2020. The majority of patients (approximately 90%) received PD-1/PD-L1 inhibitors, while the remaining 10% received combined immunotherapy (CTLA4 and PD-1/PD-L1 inhibitors). Among the 1,751 cancer patients, 259 cases that experienced high-grade irAEs (grade 3 to 5 events) were manually curated according to the National Cancer Institute (NCI) Common Terminology Criteria for Adverse Events v.5 guidelines. Additionally, algorithm-based autoimmune-like electronic health records were used to identify 339 patients who experienced any grade irAEs (referred to as all-grade irAEs). Most of these cases were grade 2 or higher events (27). The tumor tissue of these patients was sequenced using the targeted OncoPanel sequencing platform. After quality control steps, germline SNPs were imputed by utilizing ultra-low-coverage off-target reads. Then, the GWAS was conducted in the DFCI cohort to investigate the association of all variants with the time from the start of ICIs treatment to the occurrence of the two phenotypes of irAEs. For more detailed information, please refer to the original publication (27).

### Selection of instrumental variables

Several steps were followed in the selection of iVs. Firstly, for the gut microbiome, we selected SNPs associated with bacterial taxa with a p-value less than 1 × 10^−5^ for further analysis (28,29). Secondly, potential SNPs were clumped for independence in the TwoSampleMR package in R software. We used the European 1000 Genomes Project Phase 3 reference panel and set the linkage-disequilibrium threshold (r^2^) at 0.001 within a 10 Mb window size. Thirdly, we extracted SNPs from the outcome statistics and performed a harmonization procedure. SNPs that were not available in the outcome GWAS data were replaced with proxy SNPs (r^2^ > 0.8), and palindromic SNPs were removed for further MR analysis. Furthermore, F statistics of selected iVs, which indicate instrument strength, was calculated as [Beta/SE]^2^. Typically, F statistics > 10 suggest enough iVs strength to avoid weak instrument bias (30). Finally, All SNPs with positive results were re-examined using PhenoScanner package (version 1.0) in R software to investigate the presence of potential confounders. Bacterial taxa with less than 3 valid SNPs and unknown origin were excluded from the analysis to mitigate potential bias. Consequently, we included a total of 104 genus-level and 28 family-level bacterial taxa (n = 132) for further MR analysis.

### Statistical analysis

In this study, we employed several MR analysis methods to explore the potential causal relationship between gut microbiota and irAEs. The methods used included IVW, MR PRESSO, ML, weighted median, weighted mode, and a constrained maximum likelihood and model averaging based method (cML-MA-BIC). IVW and MR PRESSO were used in the primary analysis. In general, IVW provides maximum statistical power when all instruments are valid (31), while MR PRESSO identifies and removes genetic variants that deviate significantly from the variant-specific causal estimates of other variants, thereby increasing statistical power and addressing potential outliers (32). The ML method resembles the IVW approach which assumes the absence of both heterogeneity and horizontal pleiotropy. If these assumptions hold true, the ML method yields unbiased results with smaller standard errors compared to the IVW approach (33). Considering the potential existence of IV pleiotropy, we also performed pleiotropy-robust methods including weighted median, weighted mode, and cML-MA-BIC in the sensitivity analysis. These methods relax the instrumental variable assumptions. Weighted median were introduced when the exclusion restriction assumption was violated (uncorrelated pleiotropy), which typically assume fewer than 50% of genetic variants are invalid (34). The weighted-mode method clusters genetic variants based on their similarity in causal effect and estimates the overall causal effect based on the cluster with the most number of iVs (35). The cML-MA-BIC method is a novel approach developed for MR analysis, specifically addressing the issue of invalid iVs exhibiting both uncorrelated and correlated pleiotropy (violation of the independence assumption) (36). By being robust to such violations, cML-MA-BIC improves the accuracy of MR analysis, reduces Type I error, and increases statistical power (36).

Next, heterogeneity and directional pleiotropy were assessed using Cochra’’s Q statistics and MR Egger intercept. Leave-one-out (LOO) analysis was conducted to identify possible reliance on a specific variant, which involved excluding one SNP at a time for all valid SNPs in the IVW analysis. Additionally, reverse MR analysis between irAEs and the identified significant gut bacterial taxa was performed. Moreover, MVMR analysis, which could help us better understand the intricate interplay between the risk factors was further conducted (37). We considered a Bonferroni-corrected p-value of 3.89 × 10^−4^ (0.05/132) as the significance threshold for gut microbiota. Two-tailed p-value < 0.05 was considered suggestive of significance. All analyses were conducted using R packages “TwoSampleMR” (version 0.5.6), “MRPRESSO” (version 1.0), and “MRcML” (version 0.0.0.9) in R software (version 4.2.2).

## 3. Results

### 3.1 Genetic instruments and primary MR analysis

A total of 870 single nucleotide polymorphisms (SNPs) were selected as instrumental variables (iVs) for the 132 gut bacterial taxa (**Table S1**). The F statistics for each SNP ranged from 16.91 to 36.57, with a median value of 21.66. Using the IVW and MR-PRESSO methods, eight gut bacterial taxa associated with high-grade irAEs were identified with p-values < 0.05. These taxa include *Lachnospiraceae*, *Verrucomicrobiaceae*, *Ruminiclostridium6*, *Coprococcus3*, *Anaerostipes*, *Akkermansia*, *Collinsella*, and *Eubacterium (fissicatena group)*. For all-grade irAEs, seven gut bacterial taxa, including *Lachnospiraceae*, *Porphyromonadaceae*, *Roseburia*, *RuminococcaceaeUCG004*, *DefluviitaleaceaeUCG011*, *Eubacterium (brachy group)*, and *Peptococcus*, were identified. Given previous studies suggest that pre-existing autoimmune conditions such as inflammatory bowel disease, psoriasis, and rheumatoid arthritis may predispose individuals to irAEs susceptibility (38,39). We further examined the SNPs associated with the significant bacterial taxa using PhenoScanner. Only one SNP (rs11597285) for the *Collinsella* genus was found to be associated with allergic disease (e.g. allergic rhinitis and eczema) (refer to **Table S9**). However, the results of *Collinsella* remained uninfluenced after removing rs11597285 in the LOO analysis (described below). The complete results of the primary MR analysis can be found in **Table S2** and **Table S3**.

### 3.2 Main MR results and sensitivity analysis for high-grade irAEs

As shown in **Figure 2**, the IVW estimate suggested the abundance of *Lachnospiraceae* family was associated with a shortened time to high-grade irAEs (Beta = −1.22, 95% CI: −1.99 to −0.44, p = 2.17 × 10^−3^), indicating *Lachnospiraceae* serves as a risk factor for the development of high-grade irAEs. The deleterious effect remained significant in pleiotropy-robust cML-MA-BIC estimation (Beta = −1.24, 95% CI: −2.45 to −0.02, p = 4.62 × 10^−2^). Surprisingly, *Ruminiclostridium6* genus was significantly associated with an increased risk of high-grade irAEs in all MR approaches, including IVW (Beta = −2.11, 95% CI: −2.98 to −1.23, p = 2.47 × 10^−6^), cML-MA-BIC (Beta = −2.17, 95% CI: −3.49 to −0.86, p = 1.19 × 10^−3^), Weighted median (Beta = −2.43, 95% CI: −4.05 to −0.81, p = 3.29 × 10^−3^), and other methods. In addition, the IVW estimate indicated a protective effect of the *Akkermansia* genus on high-grade irAEs (Beta = 1.27, 95% CI: 0.28 to 2.25, p = 0.01), and this finding was confirmed by the IVW estimate of its paternal taxon *Verrucomicrobiaceae* (Beta = 1.27, 95% CI: 0.29 to 2.25, p = 0.01). The IVW estimate of *Anaerostipes* genus also indicated a suggestive protective effect against high-grade irAEs (Beta = 2.1, 95% CI: 0.85 to 3.35, p = 1.02 × 10^−3^), as well as cML-MA-BIC (Beta = 2.17, 95% CI: 0.57 to 3.77, p = 7.88 × 10^−3^). Moreover, significant effects of the *Coprococcus3* (Beta = −2.04, 95% CI: −2.7 to −1.39, p = 8.93 × 10^−10^), *Collinsella* (Beta = −1.12, 95% CI: −1.7 to −0.53, p = 1.7 × 10^−4^), and *Eubacterium (fissicatena group)* genus (Beta = −0.73, 95% CI: −1.01 to −0.46, p = 1.93 × 10^−7^) were all revealed by IVW estimate, indicating an increased risk of high-grade irAEs. Scatter plots reflecting the effect size of iVs on both bacterial taxa and high-grade irAEs are shown in **Figure 4**.

**Figure 2.**
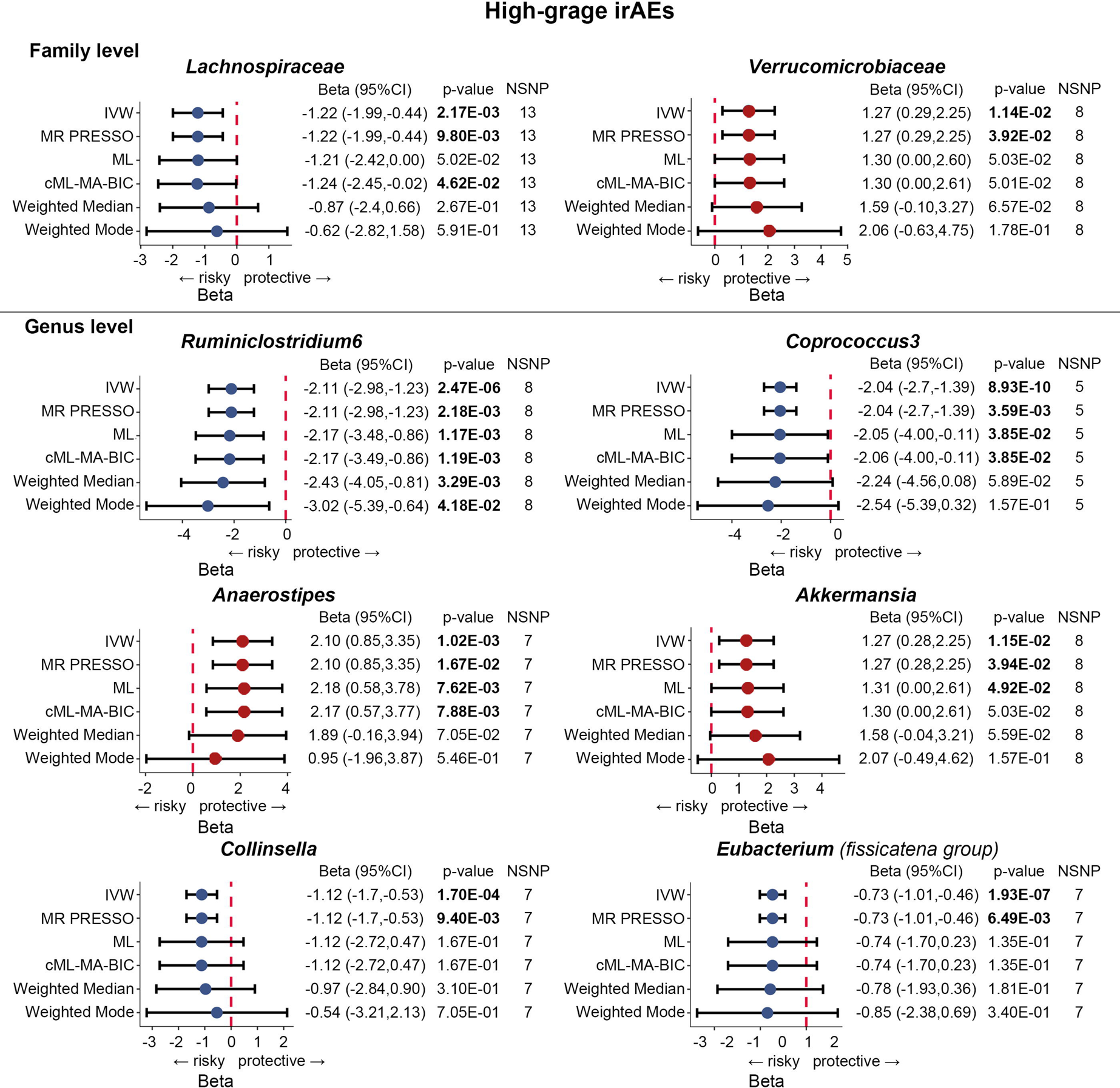
Forest plots of MR results for the causal association between the identified eight gut microbial taxa and high-grade irAEs (grade 3 to 5 events). NSNP = number of SNPs; Beta = effect size from the exposure to the outcome; CI = confidence interval.

**Figure 3.**
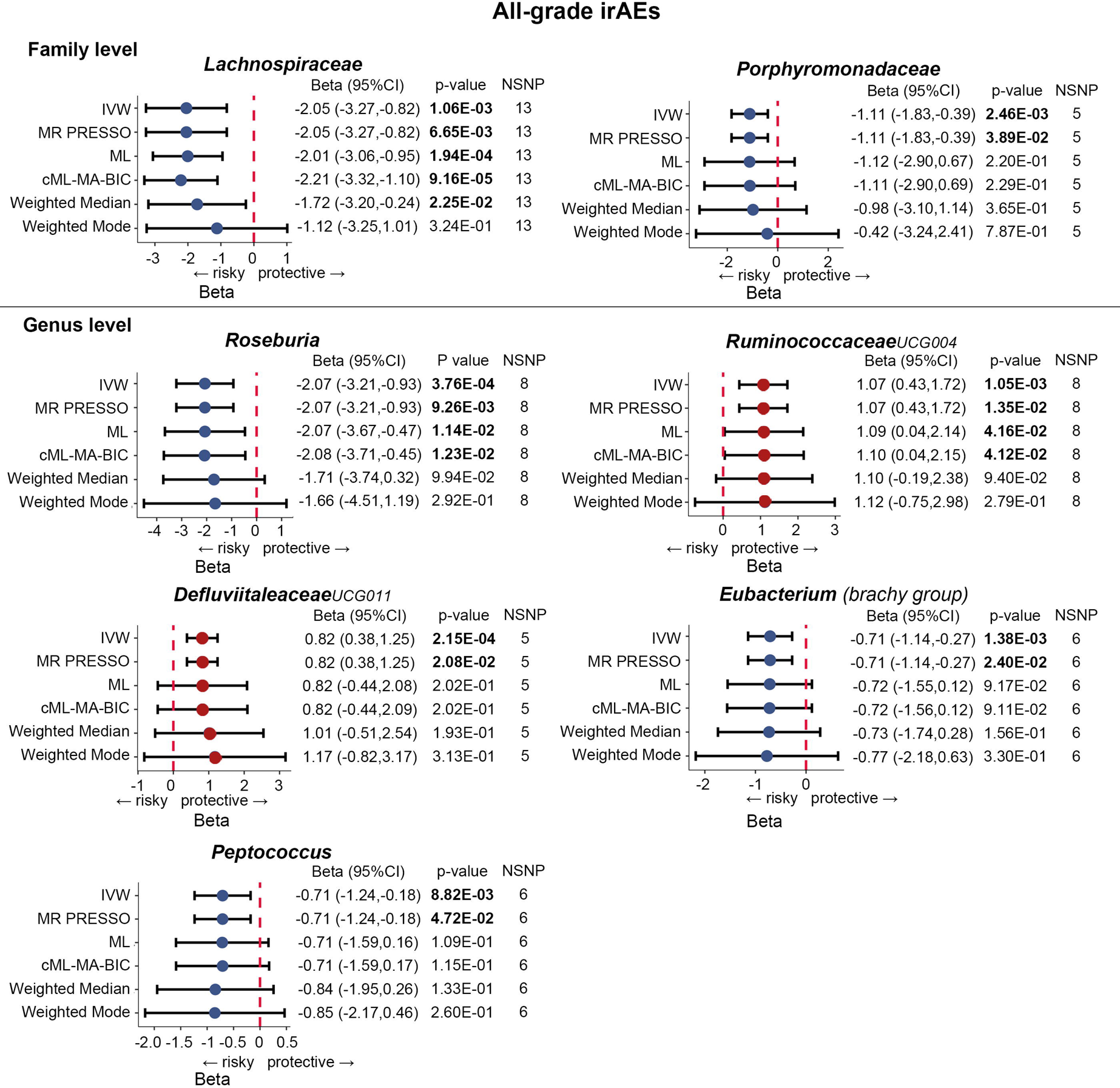
Forest plots of MR results for the causal association between the identified seven gut microbial taxa and all-grade irAEs (grade 1 to 5 events). NSNP = number of SNPs; Beta = effect size from the exposure to the outcome; CI = confidence interval.

**Figure 4.**
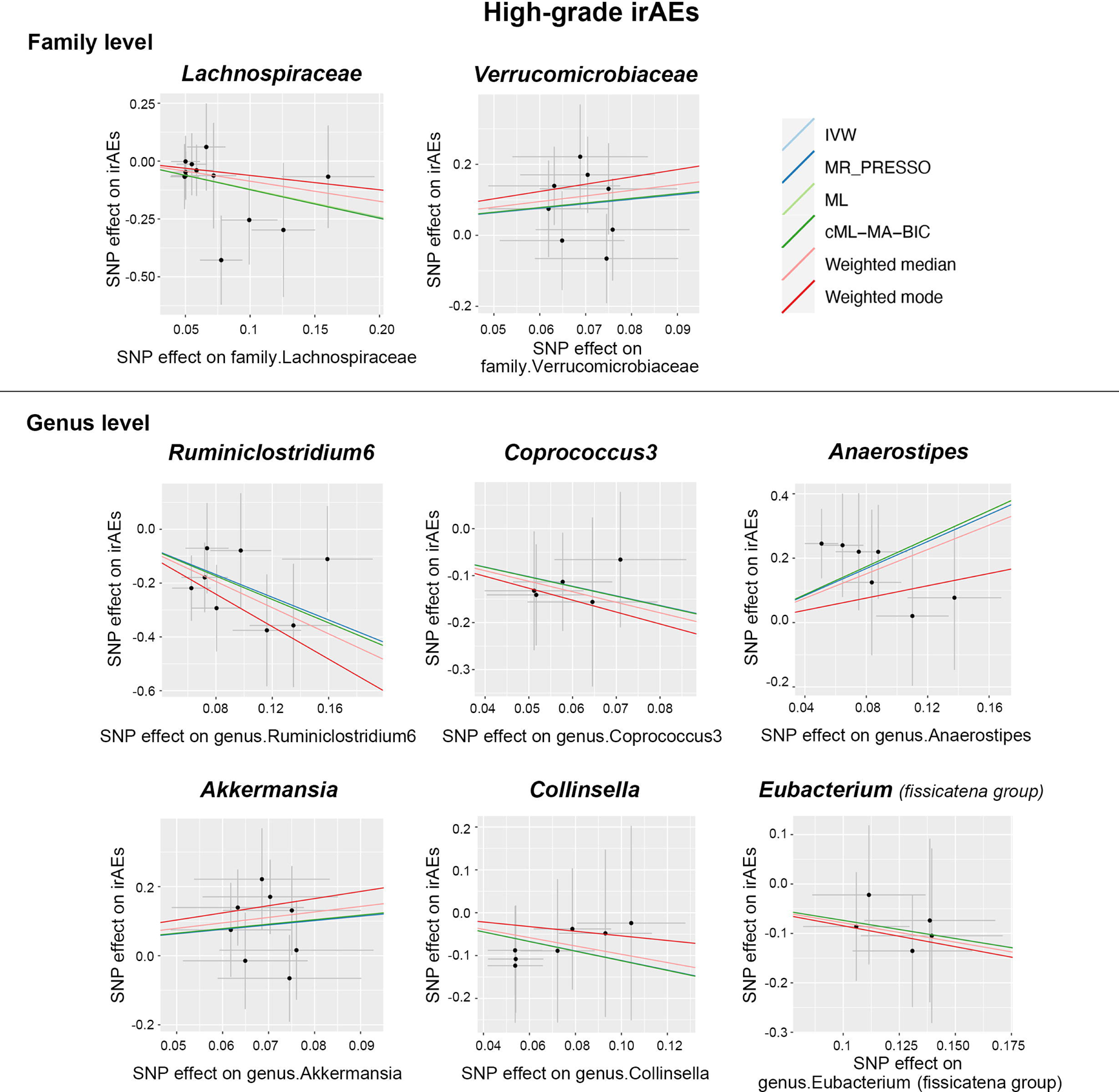
Scatter plots of MR analysis between the gut microbial taxa and high-grade irAEs (grade 3 to 5 events).

**Figure 5.**
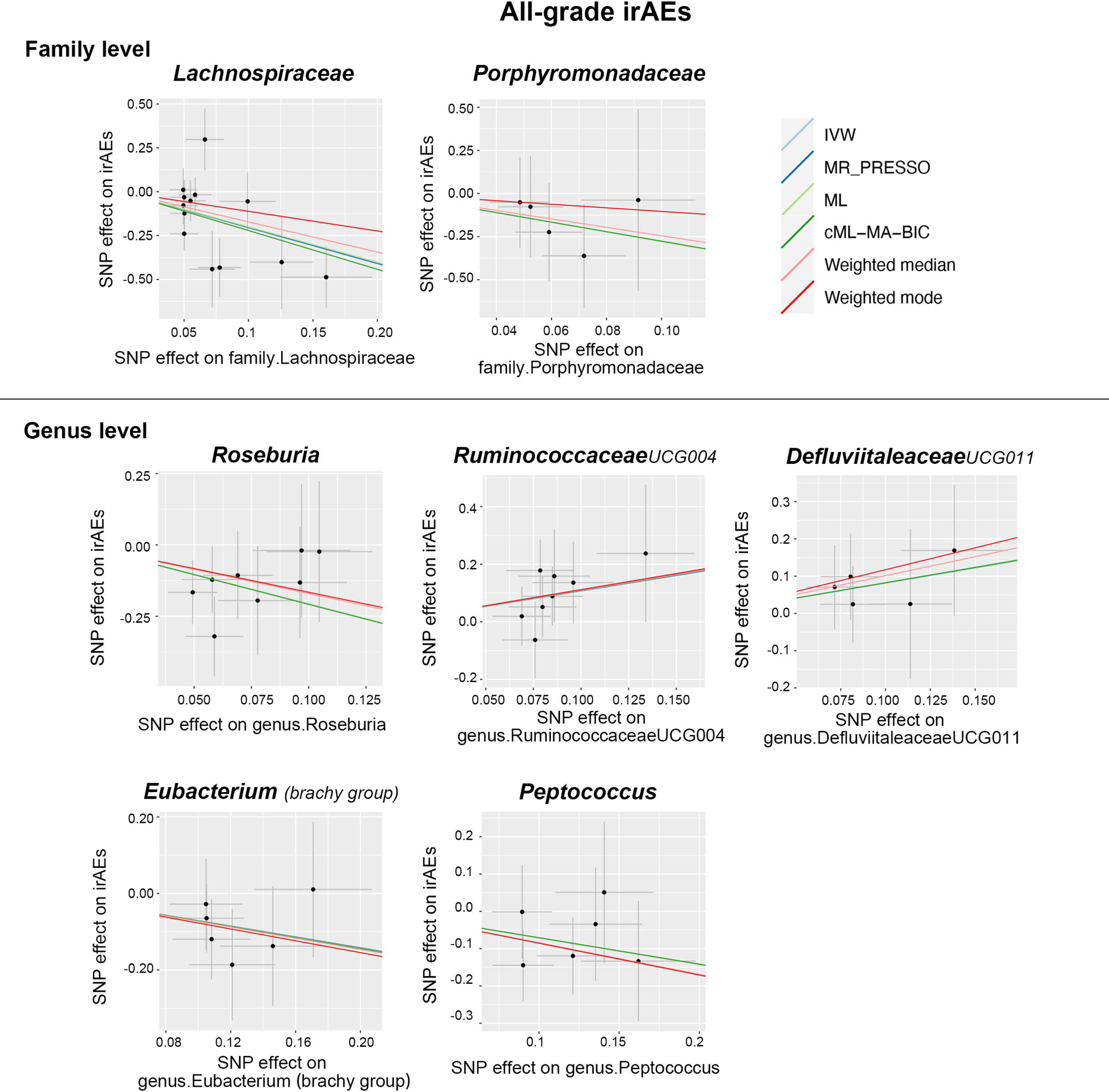
Scatter plots of MR analysis between the gut microbial taxa and all-grade irAEs (grade 1 to 5 events).

In the subsequent analysis of heterogeneity and horizontal pleiotropy, Cochra’’s Q statistics revealed no significant heterogeneity (p-value> 0.05) among the iVs for the gut bacterial taxa in high-grade irAEs analysis (see **Table 1**). No significant evidence for directional horizontal pleiotropy was found in the MR-Egger regression intercept analysis and MR PRESSO global test (**Table 1 and Table S4**). Additionally, the LOO analysis identified no predominant SNP that influenced the results (**Figure 6**). We further performed reverse MR analysis and demonstrated no reverse causation exists between high-grade irAEs and the abundance of gut bacterial taxa (**Table S6**).

**Figure 6.**
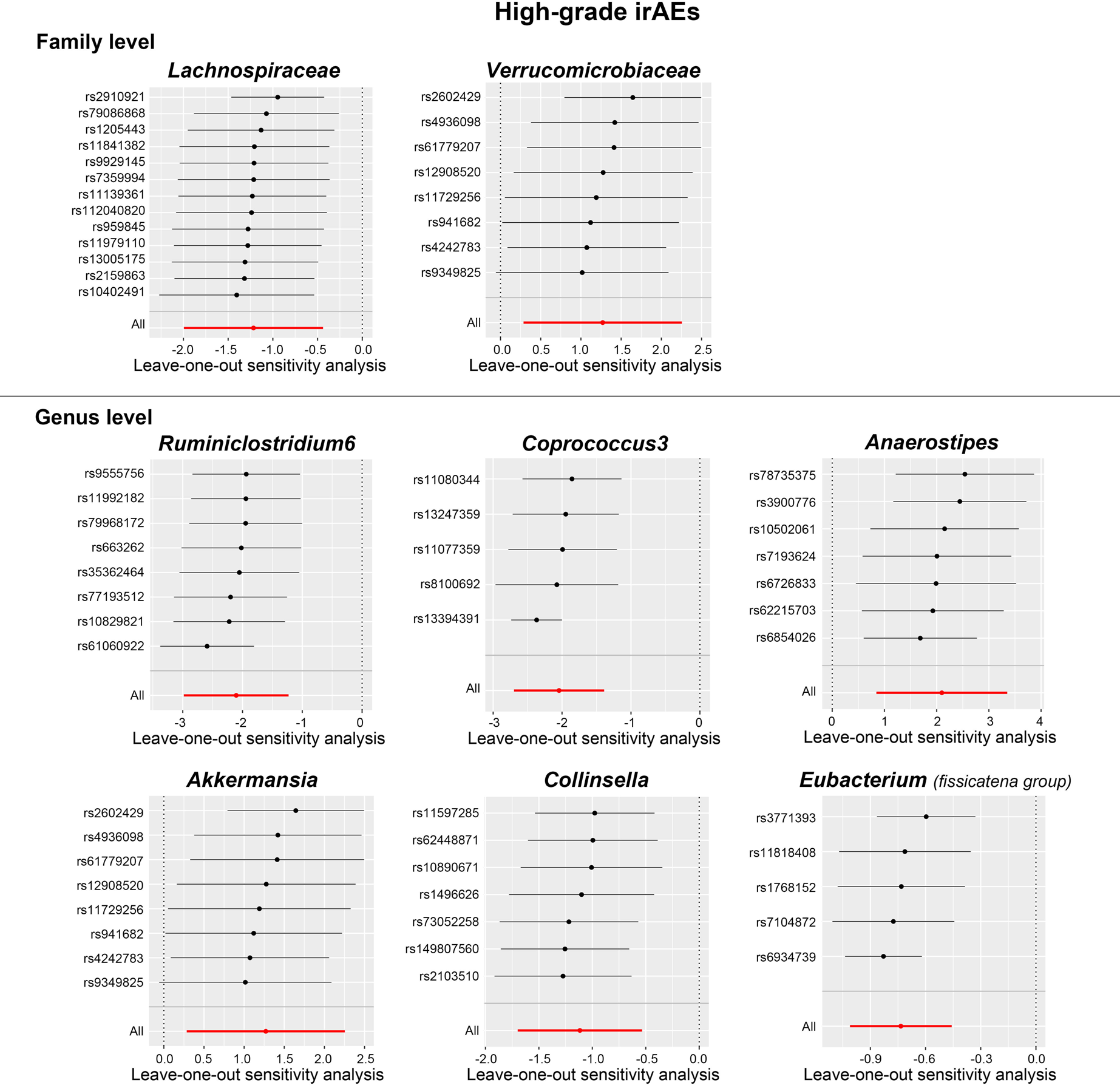
Leave-one-out plots of MR analysis between the gut microbial taxa and high-grade irAEs (grade 3 to 5 events).

**Table 1.**
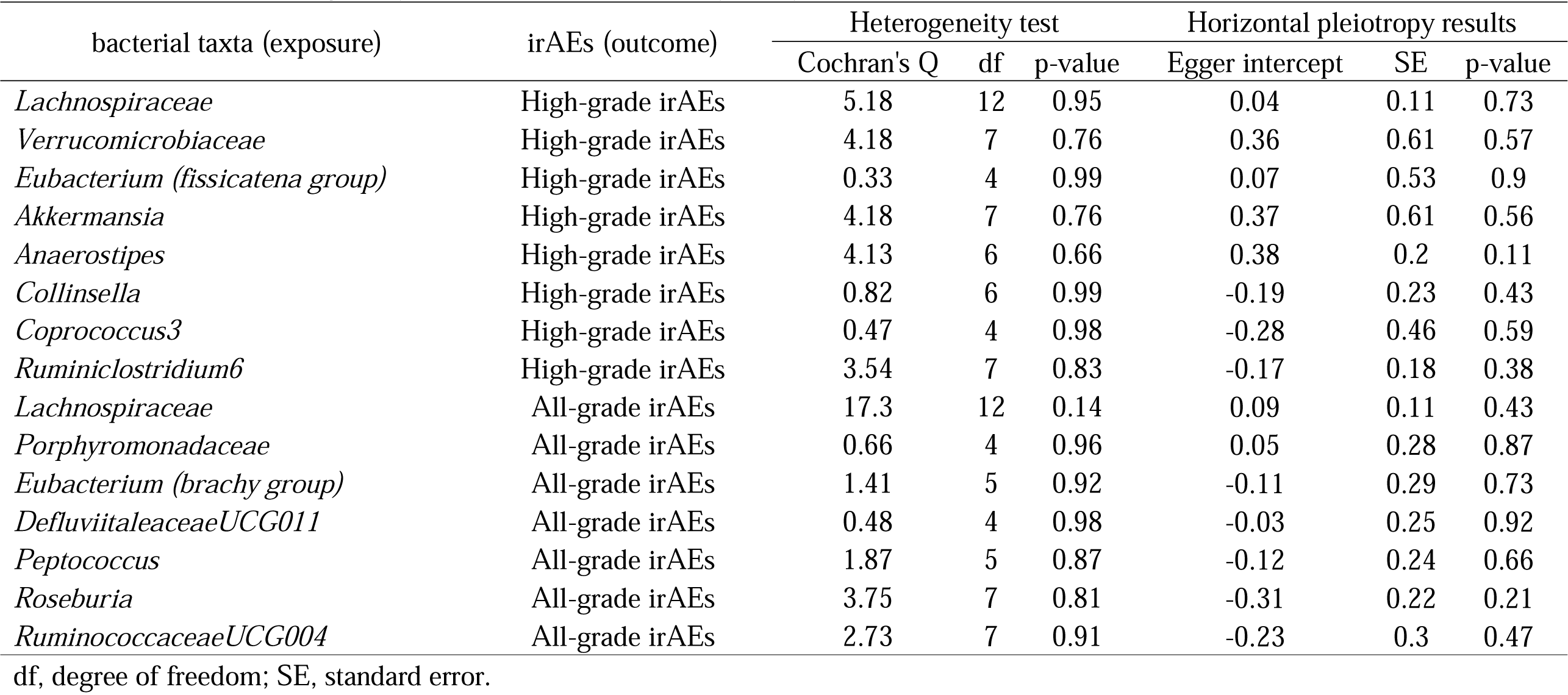
Results of heterogeneity and horizontal pleiotropy analysis.

### 3.3 Main MR results and sensitivity analysis for all-grade irAEs

Figure 3 reveals the association between bacterial taxa and all-grade irAEs. It is noteworthy that the deleterious impact of the *Lachnospiraceae* family was also detected in all-grade irAEs, as revealed by IVW (Beta = −2.05, 95% CI: −3.27 to −0.82, p = 1.06 × 10^−3^), cML-MA-BIC (Beta = −2.21, 95% CI: −3.32 to −1.1, p = 9.16 × 10^−5^), and Weighted median estimates (Beta = −1.72, 95% CI: −3.2 to −0.24, p = 2.25 × 10^−2^). The IVW estimate of *Roseburia* genus also showed an increased risk of all-grade irAEs (Beta = −2.07, 95% CI: −3.21 to −0.93, p = 3.76 × 10^−4^). Consistent results were observed in ML (Beta = −2.07, 95% CI: −3.67 to −0.47, p = 1.14 × 10^−2^) and cML-MA-BIC (Beta = −2.08, 95% CI: −3.71 to −0.45, p = 1.23 × 10^−2^) estimates. On the contrary, *RuminococcaceaeUCG004* (Beta = 1.07, 95% CI: 0.43 to 1.72, p = 1.05 × 10^−3^) and *DefluviitaleaceaeUCG011* (Beta = 0.82, 95% CI: 0.38 to 1.25, p = 2.15 × 10^−4^) were identified to decrease the risk of all-grade irAEs according to the IVW approach. Subsequently, the results for *RuminococcaceaeUCG004* were consistent with the cML-MA-BIC (Beta = 1.1, 95% CI: 0.04 to 2.15, p = 4.12 × 10^−2^) and ML method (Beta = 1.09, 95% CI: 0.04 to 2.14, p = 4.16 × 10^−2^). Moreover, the IVW estimates suggested that *Porphyromonadaceae* (Beta = −1.11, 95% CI: −1.83 to −0.39, p = 2.46 × 10^−3^), *Eubacterium (brachy group)* (Beta = −0.71, 95% CI: −1.14 to −0.27, p = 1.38 × 10^−3^), and *Peptococcus* (Beta = −0.71, 95% CI: −1.24 to −0.18, p = 8.82 × 10^−3^) may increase the risk of all-grade irAEs. Scatter plots reflecting the effect size of each IV on both bacterial taxa and all-grade irAEs are shown in Figure 5.

Similarly, Cochran’s Q statistics indicated an absence of notable heterogeneity in the iVs of gut bacterial taxa (refer to **Table 1**). In addition, the results from the MR-Egger regression intercept analysis and the MR PRESSO global test demonstrated no significant evidence of directional horizontal pleiotropy (**Table 1 and Table S4**). Next, no significant single SNP was identified in the LOO analysis that influenced the results (Figure 7). Overall, these results provide evidence for the association between specific gut bacterial taxa and the development of high-grade and all-grade irAEs, and highlight the potential role of the gut microbiota in irAEs.

**Figure 7.**
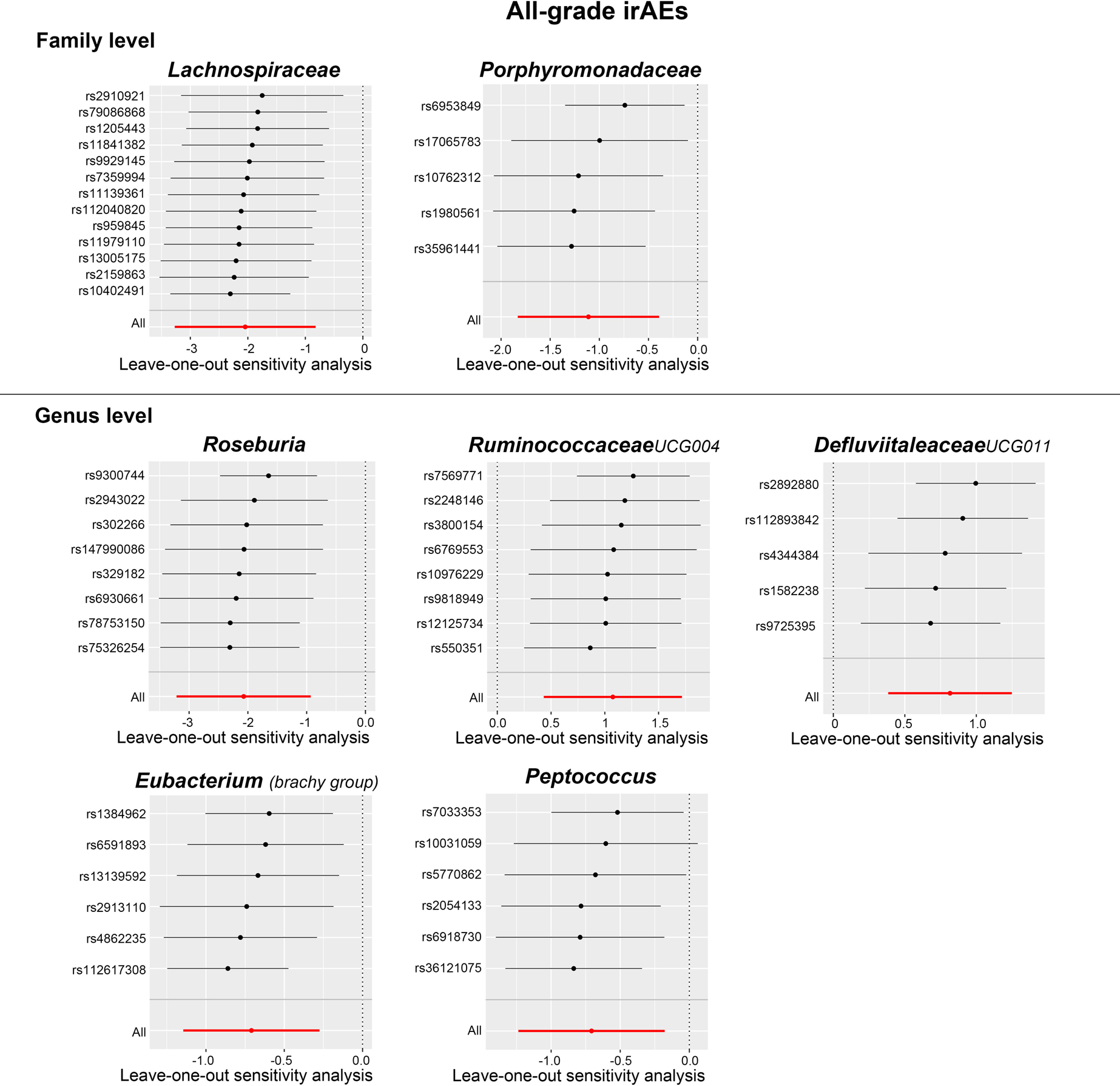
Leave-one-out plots of MR analysis between the gut microbial taxa and all-grade irAEs (grade 1 to 5 events).

### 3.4 Multivariable MR analysis

Given the strong correlation between body mass index (BMI) and gut microbiota (40,41), we incorporated BMI and weight-associated anthropometric traits, as well as several lifestyle risk factors associated with irAEs (42), into the univariable MR analysis. Detailed GWAS datasets information of these anthropometric traits and lifestyle risk factors can be found in **Table S10**. The univariable MR analysis suggested a correlation between BMI and an increased risk of all-grade irAEs (**Table 2**). Thus, we performed subsequent MVMR analysis by including BMI as an additional exposure alongside previously identified gut microbial taxa (**Table S11**). Notably, after accounting for the effect of BMI in the MVMR analysis, *Lachnospiraceae* still remained significantly associated with the risk of developing all-grade irAEs (Beta = −1.07, 95% CI: −1.96 to −0.18, p = 0.02), and high-grade irAEs (Beta = −0.91, 95% CI: −1.84 to 0.01, p = 0.05) with a marginal p value (**Table 3**).

**Table 2.**
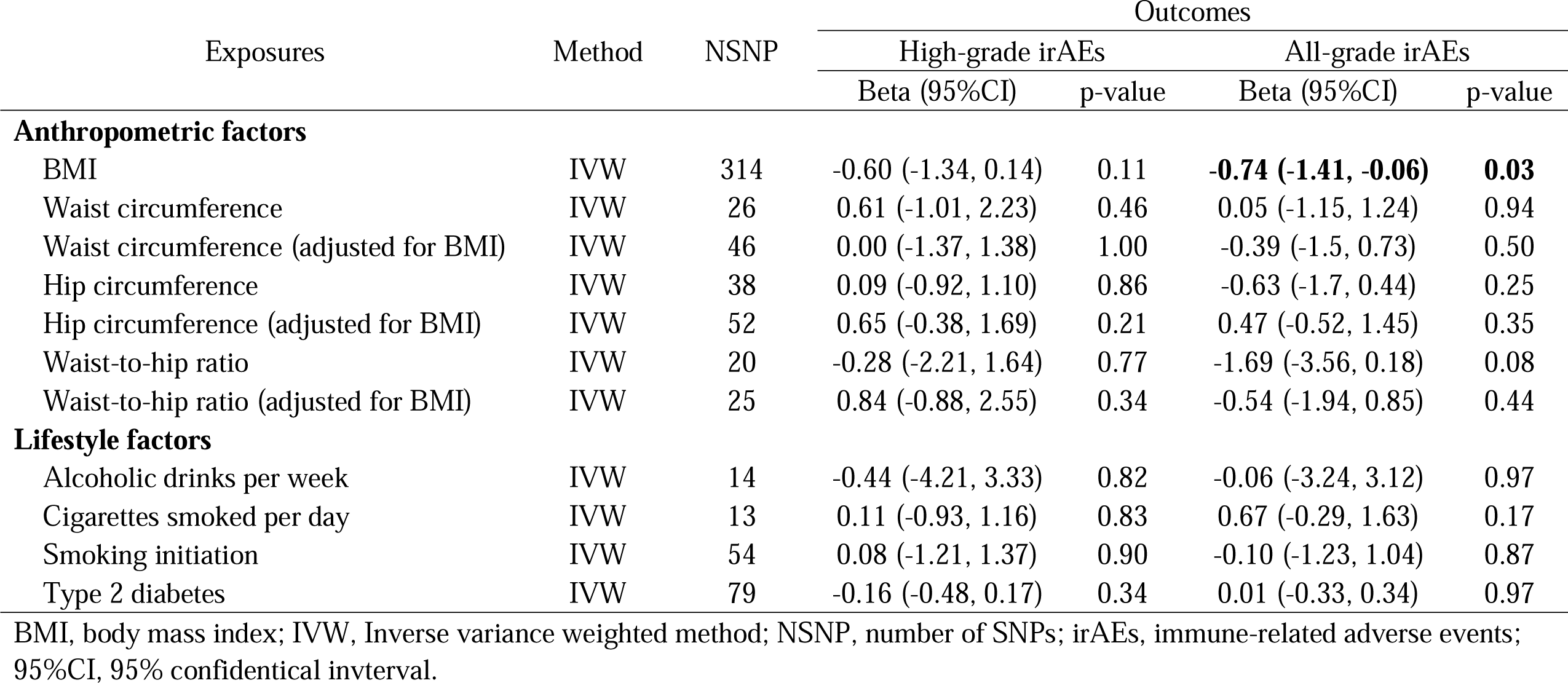
Univariate MR estimates between common risk factors and irAEs.

**Table 3.**
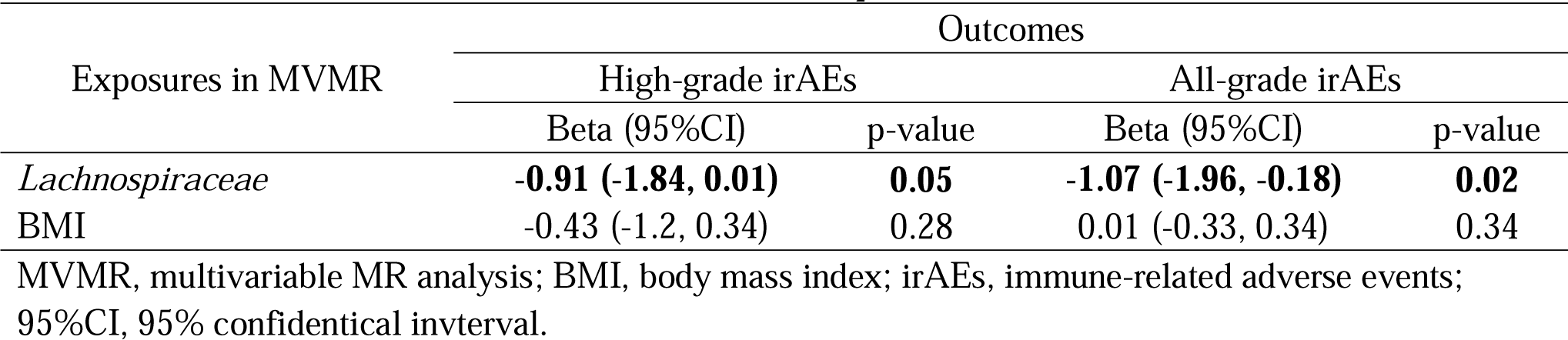
Multivariable MR estimates between *Lachnospiraceae*, BMI and irAEs.

## 4. Discussion

In this study, univariable bi-directional MR and MVMR analyses were conducted to infer the causal association between gut microbiota and irAEs. We identified fourteen gut bacterial taxa that were causally associated with high-grade and all-grade irAEs. Surprisingly, *Lachnospiraceae* strong associated with an increased risk of both irAEs phenotypes, even after accounting for the effect of BMI in the MVMR analysis. Additionally, we found robust evidence indicating that *Ruminiclostridium6* predisposes ICIs receivers to developing high-grade irAEs. *Coprococcus3*, *Collinsella*, and *Eubacterium (fissicatena group)* were also associated with an increased risk of high-grade irAEs, while *Akkermansia*, *Verrucomicrobiaceae*, and *Anaerostipes* exhibited protective roles in high-grade irAEs. For all-grade irAEs, *Porphyromonadaceae*, *Roseburia*, *Eubacterium (brachy group)*, and *Peptococcus* were associated with an elevated risk, while *RuminococcaceaeUCG004* and *DefluviitaleaceaeUCG011* were associated with a reduced risk.

Several observational studies have demonstrated associations between gut microbiota and irAEs (11,17–19,43,44). *Lachnospiraceae* species (such as *Coprococcus* and *Roseburia*), which are obligately anaerobic, variably spore-forming bacteria, were found to correlate with increased risk of various types of irAEs (17,18,45). In a more recent study, two species of the *Lachnospiraceae* family were specifically enriched in irAEs that occur in endocrine organs (46). Consistent with these findings, we also identified the *Lachnospiraceae* family, and its two genera (i.e., *Coprococcus3* and *Roseburia*) associated with an increased risk of irAEs. Importantly, the harmful influence of the *Lachnospiraceae* family was observed in high-grade irAEs and all-grade irAEs, even after adjusting the effect of BMI, which provides further validity and robustness to our study. *Ruminiclostridium6* has been less studied in irAEs, but some studies have observed its accumulation in a mouse model of DSS-induced colitis. While treated with phloretin (a dihydrochalcone flavonoid) or sodium butyrate (one of the short chain fatty acids [SCFAs]), both of which alleviates DSS-induced colitis, the abundance of *Ruminiclostridium6* was reduced (47,48). In addition, the *Ruminiclostridium* genus has also been associated with autoimmune-related diseases, such as the experimental multiple sclerosis model and Alzheimer’s disease (49,50). Based on our strong association of *Ruminiclostridium6* with increased risk of high-grade irAEs, it is suggested that *Ruminiclostridium6* may play a pivotal role in the development of autoimmune conditions and could be a potential target for relieving irAEs symptoms, although more evidence is needed.

*Akkermansia muciniphila*, an anaerobic gram-negative species that belongs to *Akkermansia* genus, and *Verrucomicrobiaceae* family, has gained much attention in immunotherapy due to their association with a favorable response in ICIs therapy (51–53). *Akkermansia muciniphila* has also been shown to exhibit a protective role in ICIs-associated colitis (54). Mechanically, Wang et al. demonstrated that *Akkermansia muciniphila* and its purified membrane protein mitigated colitis by regulating macrophages and CD8+ T cells in the colon tissue (55). Our study further supported the protective role of *Akkermansia* in high-grade irAEs. *Ruminococcaceae*, a key family of bacteria producing short-chain fatty acids (SCFAs), has been observed to be enriched in ICIs responders without severe irAEs (11). Previous studies have found that *Faecalibacterium prausnitzii*, a species belonging to the *Ruminococcaceae* family, was decreased in non-responders and those experiencing severe irAEs (11,17,45). These findings suggest a potential role of *Ruminococcaceae* as protective bacteria, possibly through the facilitation of SCFAs accumulation, in the mitigation of irAEs. *Collinsella* and *Anaerostipes* have limited evidence in irAEs, but the *Collinsella* genus has been reported to increase the production of IL-17A and enhance rheumatoid arthritis severity (56). In contrast, *Anaerostipes*, which belongs to the *Lachnospiraceae* family, was conversely associated with the risk of high-grade irAEs in our study (11,18). These discrepancies observed in previous clinical studies might be attributed to several factors, including limited sample sizes in previous observational studies, heterogeneity among the samples, and inadequate exploration of the taxonomic classification at the genus level of the gut microbiota. Therefore, a more detailed taxonomy for gut microbiota is crucial in dissecting the underlying mechanisms.

Gut microbiota plays a pivotal role in modulating human immune homeostasis, and an imbalance in gut microbial composition, known as gut dysbiosis, has been implicated in several autoimmune diseases (15,16,57). IrAEs resemble autoimmune diseases in many aspects (38,58,59). Thus, despite the underlying mechanisms by which gut microbiota manipulates the development of irAEs remain poorly understood, we hypothesize that there might be some shared etiology of microbiota in autoimmune diseases and irAEs. These mechanisms include: (1) “Molecular mimicry”: Evidence has shown that exposure to homologous amino acid sequences or epitopes of microbiota and aberrant activation of autoreactive B or T cells leads to multiple autoimmune diseases, such as multiple sclerosis (60), Guillain–Barré syndrome (61), Type 1 diabetes (62), Rheumatoid arthritis (63), and primary biliary cholangitis (64), which is referred to as “molecular mimicry” (65). It is believed that the systematic activation of the immune system during ICIs treatment triggers irAEs by bypassing self-tolerance in normal organs. One intriguing fact is that most irAEs occur in barrier organs (e.g., the intestinal tract, skin, and lungs) (58,66). This implies that the activated immune response might target the commensal microbiome as antigenic targets, although this hypothesis has not been fully demonstrated. (2) Decreased accumulation of SCFAs: SCFAs, including acetate, propionate, and butyrate, are a group of organic compounds primarily produced by the gut microbiota during the fermentation of dietary fibers. These metabolites were shown to improve the anti-cancer function of effector T cells, but they also seem to exhibit anti-inflammatory characteristics (67,68). Butyrate, one of the well-studied SCFAs, was shown to inhibit the activation of NF-κB and its downstream pathway (69), thereby reducing the production of pro-inflammatory cytokines such as IL-8 (70), while increasing the levels of anti-inflammatory factors like IL-10 (71). Moreover, SCFAs serve as a key energy source for colonocytes and maintain intestinal barrier integrity (72). Thus, the reduced abundance of SCFAs-producing bacteria along with its metabolites may participate in the development of irAEs. (3) Other mechanisms: Stimulation of the immune response by microbial-associated molecular patterns (e.g., include lipopolysaccharides (LPS), lipoproteins, flagellin and bacterial DNA) (73) and compromised vitamin B and polyamine metabolism that associated with gut dysbiosis (19) may also contribute to irAEs.

Taken together, the gut microbiota and the human immune system maintain a delicate balance under normal physiological conditions. Once the balance has been disturbed (e.g., ICIs treatment), the dysregulated microbiota might lead to the development of undesirable irAEs. The primary management strategy for irAEs (> grade 2) involves the suspension of ICIs and/or utilizing immunosuppressive therapy (74). Nevertheless, one concern is that discontinuing ICIs or using immunosuppressants may compromise treatment efficacy. Ideally, approaches to boost ICIs efficacy while reducing the accompanied irAEs are to be expected in the future. FMT, an approach to modulate gut microbiota, has been shown to increase ICIs efficacy in melanoma patients (75,76), and emerging evidence has demonstrated the mitigation of ICIs-related colitis through FMT in clinical practice (77). Interestingly, while irAEs and ICIs efficacy are often considered two sides of the same coin, certain gut bacteria, such as *Akkermansia muciniphila* and *Faecalibacterium prausnitzii*, have been shown to ameliorate irAEs and reinforce ICIs efficacy at the same time (78). This suggests that targeting gut microbiota could be an ideal approach to relieve irAEs symptoms and maintain ICIs efficacy, but further real-world evidence is needed to support this hypothesis.

Our study possesses several strengths. Firstly, we applied the MR approach to infer the causal associations between gut microbiota and irAEs, which effectively mitigates the influence of confounding factors and provides robust causal inference. Secondly, we conducted reverse MR analyses, confirming the absence of reverse causation, thereby enhancing the validity of our study. Thirdly, we applied MVMR in our analysis which further strengthened the validity and robustness of our study. Moreover, we incorporated several pleiotropy-robust methods such as MR PRESSO and cML-MA-BIC, further strengthening the robustness of our study. However, there are also certain inherent limitations in our study that should be considered when interpreting the findings. Firstly, our analysis is based on European-derived GWAS summary statistics, which might confine the generalization of the findings to other populations. Secondly, due to the utilization of summary statistics instead of raw data in the analysis, subgroup analyses based on ICIs regimes (e.g. PD-1/PD-L1 group, CTLA-4 group, and combined therapy group) could not be performed. Thirdly, the gut microbiota is shaped by multiple environmental factors, which confines the number of the identified significant gene loci in the GWAS (26). Thus, we relaxed the significant threshold of iVs to 1×10^−5^ (29) and employed Bonferroni correction to mitigate potential false positive results.

## 5. Conclusion

In conclusion, our univariable and multivariable MR analysis identified a strong causal association between *Lachnospiraceae* and irAEs, along with several other gut microbial taxa, such as *Akkermansia* and *Ruminiclostridium6* etc.. However, whether the FMT or probiotics could be used as interventional approaches to mitigate irAEs while reserving ICIs efficacy, additional randomized clinical trials (RCTs) are warranted. Furthermore, in-depth investigations are needed to elucidate the precise mechanisms through which the gut microbiota influences the development of irAEs.

## Declarations

## Data Availability

The GWAS summary statistics for gut microbiota are available at www.mibiogen.org, and GWAS summary statistics for irAEs are available at https://zenodo.org/record/6800429.

## Acknowledgment

We acknowledge all the investigators who have made their GWAS data available.

## Author contributions

B. L. conceptualized this study. B. L., Z. L., and T. J. were involved in the analyses and manuscript drafting of this study. X. G. and X. Z. were involved in the data curation. B. L., T. J. X. Y., Z. C., and L. D. were involved in the interpretation of the methodology and results. B. L. and Z. L. were involved in the visualization of the results. B. Z. was involved in obtaining funding and critical revision of the manuscript.

## Funding

This study was supported by the National Nature Science Foundations of China (Grant number 82203108), China Postdoctoral Science Foundation (Grant number 2022M722275), and the Key R&D Program of Sichuan Province, China (Grant number 2023YFS0278).

## Conflict of Interest

All authors declare no conflict of interest.

## Supplementary Information

**Table S1.** All Instrumental variables used for gut microbiota in the MR analysis.

**Table S2.** Full results of primary MR analysis between bacterial taxa and all-grade irAEs.

**Table S3.** Full results of primary MR analysis between bacterial taxa and high-grade irAEs.

**Table S4.** Results of ΜR PRESS Global test.

**Table S5.** Instrumental variables for irAEs in the reverse MR analysis.

**Table S6.** Results of reverse MR analysis between all-grade irAEs and the identified significant gut bacterial taxa.

**Table S7.** Results of reverse MR analysis between high-grade irAEs and the identified significant gut bacterial taxa.

**Table S8.** Results of heterogeneity and horizontal pleiotropy analysis in reverse MR analysis.

**Table S9.** PhenoScanner results of instrumental variables used for significant bacterial taxa in the MR analysis.

**Table S10.** Description of datasets used for weight-associated anthropometric and lifestyle factors in univariable MR analysis.

**Table S11.** Multivariate MR estimates between gut microbiota (adjusted for BMI) and irAEs.

---

**Figure S1.**
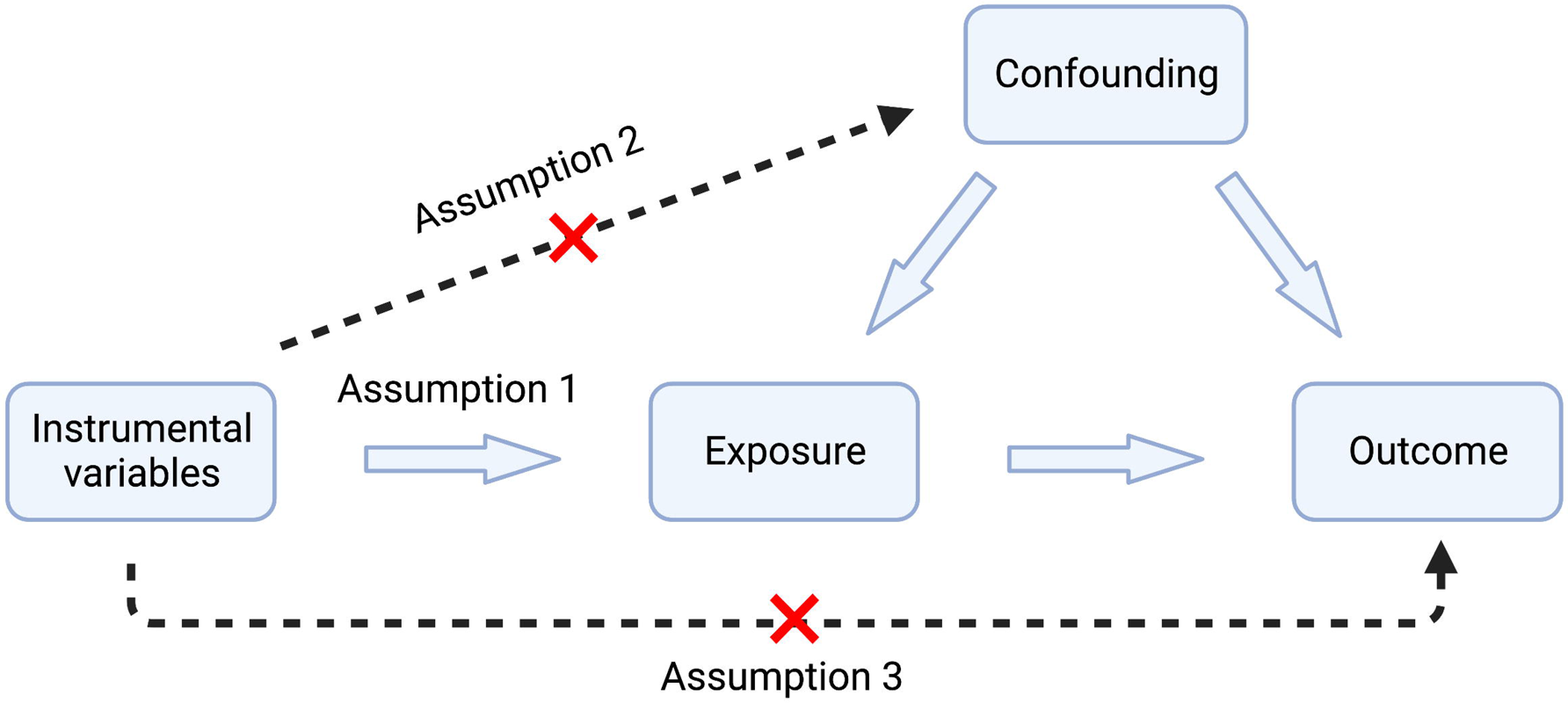
Illustration of MR assumptions

## Notes

### Competing Interest Statement

The authors have declared no competing interest.

### Author Declarations

This study was conducted using public-available data. The GWAS summary statistics for gut microbiota are available at www.mibiogen.org, and GWAS summary statistics for irAEs are available at https://zenodo.org/record/6800429.

### Summary of Updates

We added multivariable MR analysis in this version.

